# Factors Associated with False Positive Predictions in a Logistic Regression Model of High-Intensity Mental Health Service Use

**DOI:** 10.64898/2026.01.03.26343376

**Authors:** Bharadwaj V. Chada, Robert Stewart

## Abstract

**Background:** Predictive models in mental health can identify service users at risk of high-intensity care, enabling proactive interventions. However, false positive predictions may lead to over-medicalisation, inequitable resource use, and stigma. Understanding the factors associated with false positives can improve model interpretation and real-world application.

**Aims:** To evaluate a previously validated prediction model for high-intensity (top decile) service use and identify false positive (FP) vs. true positive (TP) predictions, examining factors across three timeframes: baseline (e.g. demographics and referral information), the prediction period (W1; the first three months after initial mental health service assessment), and the immediate (three month) period following prediction (W2).

**Methods:** Of mental health services users assessed between 2007-2024, 4,174 TP and 17,644 FP predictions were compared. Evaluated covariates included demographics, referral source, service use, diagnoses, medications, and recorded symptoms. Logistic regression was used to identify associations with FP outcomes across the three time periods.

**Results:** At baseline, FP predictions were more common among Asian service users, those living with family, and users referred by their GP, whilst TP predictions were associated with voluntary or probation service referrals. During W1, FP predictions were associated with higher community treatment days, crisis attendances, and service users managed in the outpatient setting. TP predictions were associated with antipsychotic use and engagement with multiple care teams. In W2, TP predictions were more likely among service users who remained outpatients, had higher inpatient days, or received multidisciplinary care, and FP more likely among those with substance use, multiple address changes, or ongoing crisis attendances.

**Conclusion:** The approach here to predictive modelling highlights the importance of considering features (here at baseline and W1) which may influence a model’s accuracy at the point the prediction is communicated, and those subsequent events (W2) which might indicate targets for intervention to prevent the adverse outcome. Both need to be incorporated in clinical interface communications if models are deployed in practice.

## Introduction

Predictive modelling in mental healthcare enables health systems to learn from past data to improve care delivery and outcomes. Models developed from routinely collected electronic health record (EHR) data can be used for a range of purposes, including accurately identifying high-intensity service users, implementing proactive and personalised interventions, and allocating resources efficiently^1^. Predictive tooling in mental health has nonetheless languished behind other medical specialties, due to a lack of objective biomarkers and a consequent reliance on self-reported symptoms and clinician judgement, limitations in accessing and analysing mental health datasets, and concerns surrounding ethics and interpretability^2^. Moreover, predictive models for mental health are often imperfect, and risk perpetuating demographic biases. False positive predictions may contribute to over-medicalisation, and, crucially, risk stigmatising already vulnerable groups of service users, whilst false negative predictions may lead to delays in treatment initiation and adverse clinical outcomes^3^.

Accordingly, even high-performing predictive models warrant detailed interrogation. Beyond improving interpretability and clinician confidence, this model evaluation can help to identify factors that influence predictive accuracy and uncertainty, enabling transparent communication of risk. Furthermore, there is clinical value in determining post-prediction circumstances that affect outcomes, thereby providing evidence to guide ethical and actionable use of model outputs.

The current project extends earlier work, in which a logistic regression model used EHR data from the first 3-months after initial assessment by mental health services, to predict high-intensity service use over the following 12 months^4,5^. The model performed strongly, achieving AUCs of ∼0.8 across development and validation datasets, with key predictors including high levels of service use during the first three months, schizophrenia and eating disorder diagnoses, and service users living alone.

The aim of the project presented here was to evaluate this model further, focussing specifically on patients predicted to have high intensity service use, and to compare those who did or did not have this outcome at the end of the twelve-month observation period. The project sought to ascertain the factors that may be more or less suggestive of truly being a high-intensity service user. The model’s predictions may be more credible when they are supported by factors that are strongly associated with high-intensity service use. Conversely, clinicians may exercise more caution when predictions rely on factors that are weakly associated with the outcome.

## Methods

### Setting

This analysis was conducted using data from the South London and Maudsley NHS Foundation Trust (SLaM), a specialist mental health care provider serving a catchment population of approximately 1.3 million residents across the London boroughs of Lambeth, Southwark, Lewisham, and Croydon. Data were accessed via SLaM’s Clinical Record Interactive Search (CRIS) platform, which enables the extraction of structured and unstructured (using natural language processing (NLP) tools) anonymised patient data from electronic health records (EHRs). This project was conducted in partnership with the NIHR Maudsley Biomedical Research Centre (BRC) and the Institute of Psychiatry, Psychology, and Neuroscience (IoPPN) at King’s College London. All analyses were performed using Python 3.11.7 on Jupyter Notebook 7.0.8.

### Recap of Predictive Model Development & Validation

The dataset used for this analysis was the same as that employed in the initial model development and validation study, comprising 46,183 patients assessed between 2007 and 2024^4^. In the earlier model, the index date was defined as 3-months following a patient’s first clinical assessment within SLaM. Individuals were eligible for inclusion if they were aged 18 years or older at the time of assessment, and had an accepted referral to SLaM. Predictor variables were derived from routinely collected data during the initial 3-month period of care, and these were used to estimate the likelihood of high-intensity service utilisation over the subsequent 12 months.

During the model development phase (2007–2011), discrimination was strong, with an AUROC of 0.79. At the selected operating threshold, the model achieved a sensitivity of 0.82 and a specificity of 0.54. When applied to later cohorts, including 2012–2017 and 2018–2023, the model maintained good performance. Across all validation periods, including those spanning the COVID-19 pandemic, discrimination remained stable (AUROC 0.79–0.83) with consistently high sensitivity.

### False Positive Analysis

The logistic regression model was applied to the full patient cohort, and predictions were compared with actual outcomes over the subsequent 12 months. The evaluation presented here focused only on cases predicted to be at risk of high-intensity service use according to the algorithm derived from this model. Each of these cases was assigned a classification flag: true positive (TP; predicted high-intensity use, and confirmed as such), or false positive (FP; incorrectly predicted to be a high-intensity user). Among 21,818 patients predicted to be positive, 4,174 were classified as TP and 17,644 as FP. For each patient, the original predictor variables were retained and supplemented with additional variables spanning several domains: demographics, service use, professionals involved, socioeconomic factors, physical health diagnoses, mental health diagnoses, medications, symptoms, substance misuse, as well as suicide, self-harm and other such risk assessments.

Prediction modifiers were further categorised by timing: baseline (information that would have been available at the time of initial referral, including the source of the referral and demographic characteristics), W1 (where ‘W’ denotes ‘window’; features occurring within the initial 3-month period after first assessment, i.e. up to the point when the prediction was made), and W2 (events occurring in the immediate 3-months post-prediction). This structure allowed examination of factors present both at, and shortly after, prediction that may influence whether an initial positive prediction was incorrect. The resulting baseline dataset comprised this longlist of predictors alongside the binary outcome of interest (FP vs. TP predictions) for further analysis.

### Model development

Within each prediction modifier category, variables with high levels of missing data (>10%) were excluded from further consideration. Univariate analyses were then conducted to identify modifiers potentially associated with the outcome (p < 0.10) within their respective categories.

This p-value was chosen to ensure that potentially informative modifiers were not prematurely excluded, as these variables could later be combined into composite predictors during clinical translation. The remaining variables were combined into an initial multivariable logistic regression model. To reduce multicollinearity, pairs of highly correlated variables (variance inflation factor > 0.8) were identified and one of each pair removed. The model was then re-estimated using the remaining modifiers. Only variables that retained statistical significance at the p < 0.10 level in this combined model were retained for the final modifier set, and these are listed in Table 1. Model performance was subsequently evaluated using this final set of variables.

**Table 1:** Final feature set included in the logistic regression model, grouped by category. W1 and W2 indicate the time window during which each prediction modifier was recorded (W1: initial three months from first assessment; W2: subsequent three months post-prediction).

| Demographics & Referral Information | Service Usage | Professionals Involved | Mental Health | Medications | Substance Misuse | Socioeconomic Factors | Other risk assessments | Symptoms |
| --- | --- | --- | --- | --- | --- | --- | --- | --- |
| Ethnicity - Asian or Asian British (Bangladeshi) | Accepted Team Count (W1) | Care Coordinator Count (W1) | Personality Disorder Diagnosis (W2) | Anticonvulsant Usage (W2) | Cannabis Usage (W2) | Address Change (W1) | Brief Risk Assessment Screening Result - 'Need for Further Investigation/ Referral' (W1) | Anxiety (W2) |
| Ethnicity - Black or Black British | Accepted Team Count (W2) | Clinical Psychology Event Count (W2) |  | Antidepressant Usage (W2) | Cocaine Usage (W2) | Address Change (W2) | Brief Risk Assessment Screening Result - 'Not Recorded' (W1) | Derailment of Speech (W2) |
| Ethnicity - Mixed Race (Other) | Crisis Attendances (W1) | Consultant Event Count (W2) |  | Distinct Generic Medications (W2) |  | Temporary Address Count (W2) | Brief Risk Assessment Screening Result - 'Significant Risk' (W1) | Disorganised Symptoms (W1) |
| First Assessment Source - ICD 10 | Crisis Attendances (W2) | Consultant Specialty – Forensic Psychiatry (W2) |  | Lithium Usage (W2) |  |  | Other Alert Recorded (W2) | Early Morning Waking (W1) |
| Lives With Status - Other | Days in Contact with Home Treatment Team (W1) | GP Recorded (W1) |  | Oral atypical antipsychotics (W1) |  |  |  | Guilt (W2) |
| Lives with Status - Other Relatives | Inpatient Days (W2) | HCA Event Count (W2) |  | Typical antipsychotics - long-acting injectables (W1) |  |  |  | Mood Instability (W1) |
| Lives With Status - Partners and Children | Outpatient Status (W1) | Nurse Event Count (W1) |  | Typical antipsychotics - long-acting injectables (W2) |  |  |  | Poor Appetite (W2) |
| Marital Status - Not Known | Outpatient Status (W2) | OT Event Count (W2) |  |  |  |  |  | Thought Withdrawal (W2) |
| Marital Status - Widowed/ Surviving Civil Partner | Total Document Count (W2) | Social Worker Event Count (W2) |  |  |  |  |  | Weight Loss (W2) |
| Referral Source - Carer |  |  |  |  |  |  |  |  |
| Referral Source - Other |  |  |  |  |  |  |  |  |
| Referral Source - Other Clinical Specialty |  |  |  |  |  |  |  |  |
| Referral Source - Probation Service |  |  |  |  |  |  |  |  |
| Referral Source - Voluntary Sector |  |  |  |  |  |  |  |  |
| Referral Status - Discharged |  |  |  |  |  |  |  |  |

## Results

Table 2 compares key demographic, clinical, and referral characteristics between TP and FP predictions. TP predictions were associated with higher service use during W1 (mean 24.9 inpatient days for TP vs. 3.3 for FP), more community contact days, and current inpatient care at the time of prediction (32.3% vs. 0.5%). Members of this group were also more frequently referred from an emergency department (ED) and had a higher prevalence of schizophrenia-related diagnoses, whereas those with FP predictions were more likely to be GP referrals and outpatients at the time of prediction. Black or Black British patients were more represented among TP (24.0% vs 16.7%), while White patients were more common among FP (68.2% vs 62.5%).

**Table 2:**
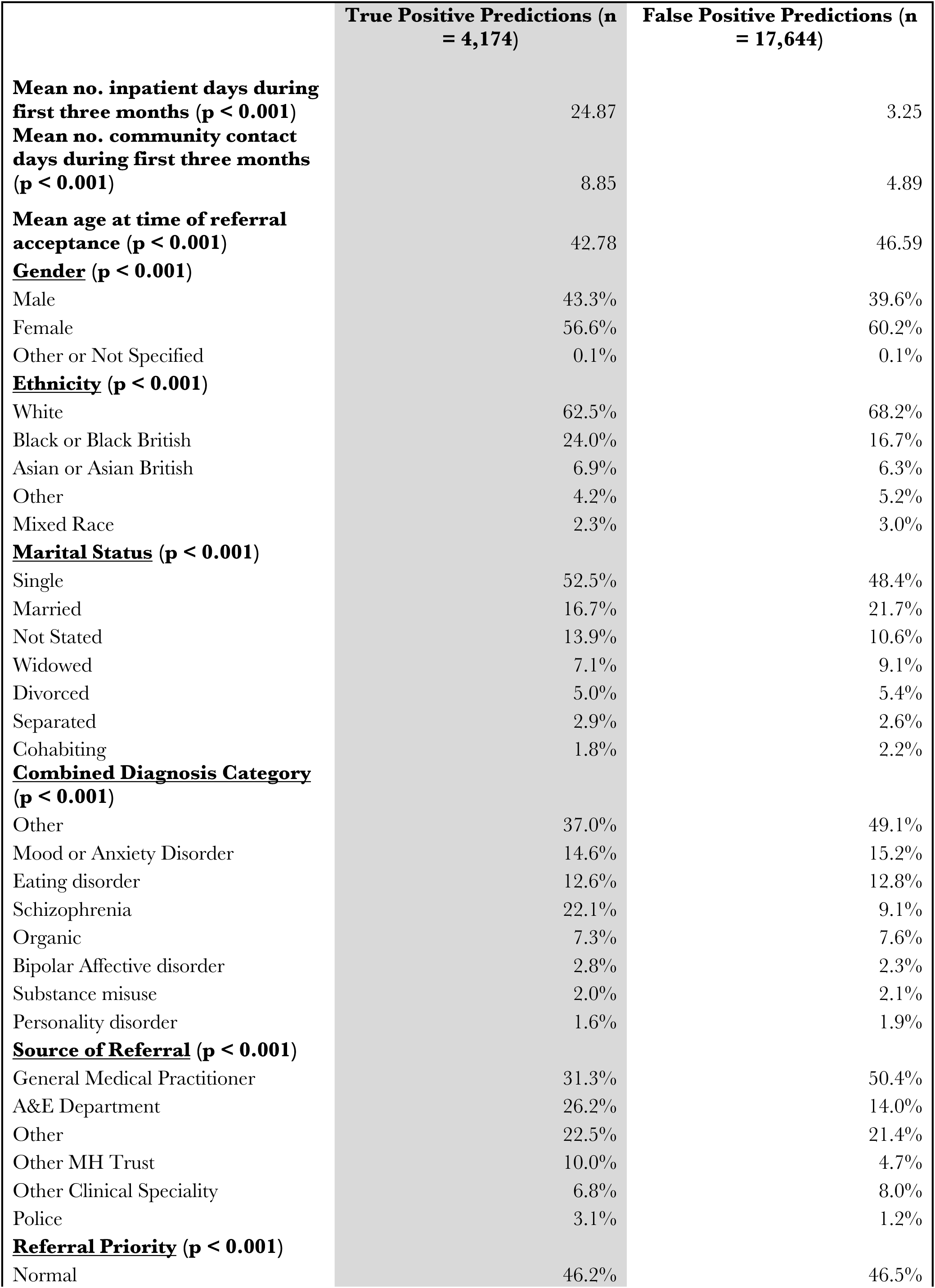
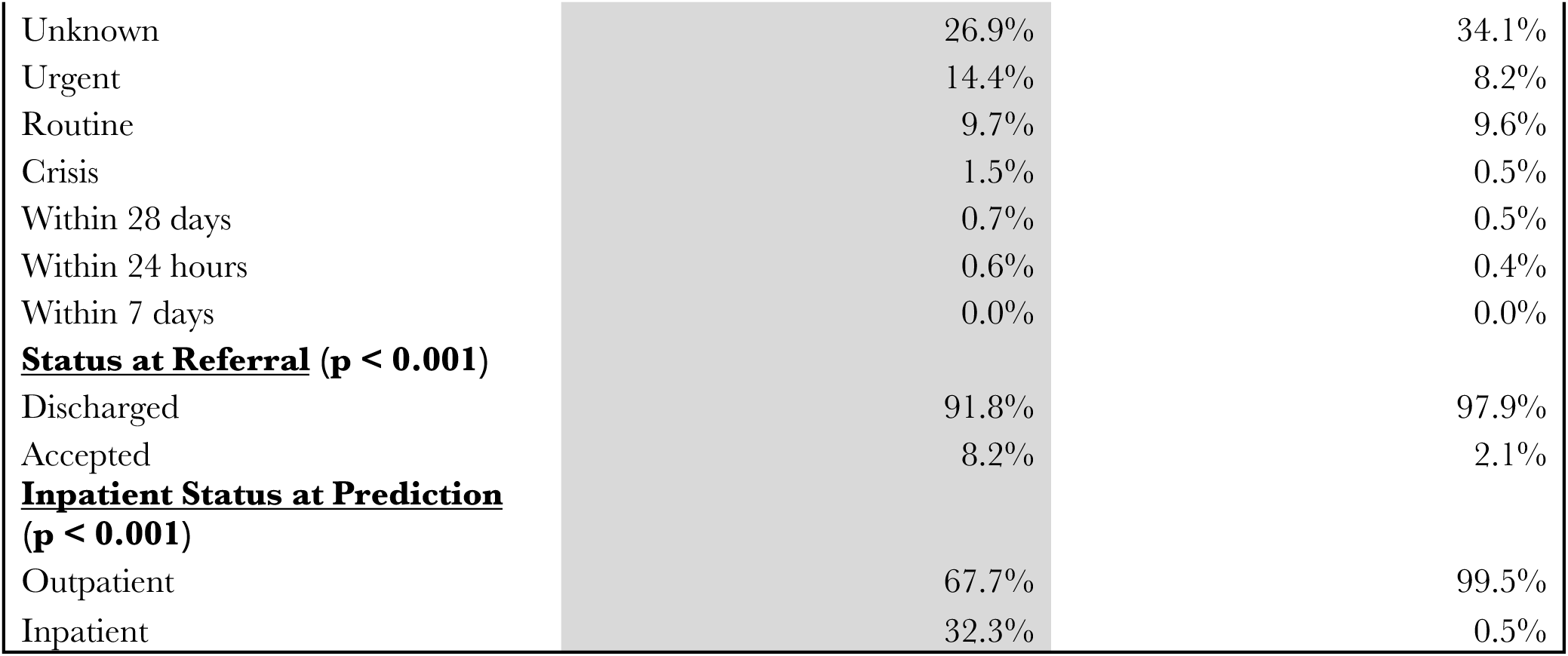
Comparison of demographic, clinical, and referral characteristics between true positive and false positive predictions.

The model was evaluated using a precision–recall framework, and a 0.18 threshold was chosen to favour precision. This prioritisation helps ensure that predictions identified as high-risk are reliable for clinical consideration, which is particularly important when decisions may be made based on these outputs. This focus on precision was intentional: by maximising confidence in the model’s positive predictions, it is possible to identify service users most likely to require high-intensity care and intervene proactively. Overall, the prediction modifier model demonstrated strong discriminative ability (AUROC 0.89), although this is skewed by the high prevalence of false positive service users within the dataset.

### Prediction modifiers

A range of modifiers at baseline, the initial 3-month window from first assessment to original model prediction (W1), and the immediate 3-month window post-prediction (W2) were evaluated to understand factors associated with the accuracy of the model’s prediction of high-intensity service users.

### Baseline

Baseline modifiers include demographic characteristics, referral information, and initial assessment details. Table 3 presents logistic regression coefficients, p-values, and 90% confidence intervals for factors associated with FP predictions. Age and gender were included in the model but were not significantly associated with FP predictions (all p > 0.4). Negative coefficients indicate factors associated with a lower likelihood of a FP outcome, while positive coefficients indicate factors associated with a higher likelihood.

**Table 3:** logistic regression results showing baseline features associated with a false positive prediction, with corresponding coefficients, p-values, and 90% confidence intervals.

| **<u>Variable</u>** | **<u>Coefficient</u>** | **<u>P value</u>** | **<u>90% confidence interval</u>** |  |
| --- | --- | --- | --- | --- |
| **First Assessment Source - ICD 10** | -0.7186 | 0.0000 | -1.0168 | -0.4204 |
| **Referral Source - Voluntary Sector** | -1.8790 | 0.0228 | -3.4963 | -0.2617 |
| **Ethnicity - Mixed Race (Other)** | -0.6730 | 0.0255 | -1.2636 | -0.0824 |
| **Ethnicity - Black or Black British** | -0.2799 | 0.0274 | -0.5287 | -0.0312 |
| **Referral Source - Other** | -0.2958 | 0.0364 | -0.5729 | -0.0187 |
| **Referral Source - Probation Service** | -1.2587 | 0.0383 | -2.4497 | -0.0677 |
| **Referral Source - Carer** | -1.5387 | 0.0425 | -3.0253 | -0.0521 |
| **Referral Status - Discharged** | 1.1556 | 0.0000 | 0.8881 | 1.4231 |
| **Lives with Status - Other Relatives** | 0.4813 | 0.0193 | 0.0782 | 0.8845 |
| **Lives With Status - Partners and Children** | 0.4720 | 0.0206 | 0.0725 | 0.8715 |
| **Marital Status - Widowed/Surviving Civil Partner** | 0.6547 | 0.0284 | 0.0692 | 1.2401 |
| **Lives With Status - Other** | 0.3159 | 0.0420 | 0.0114 | 0.6205 |
| **Ethnicity - Asian or Asian British (Bangladeshi)** | 0.9458 | 0.0705 | -0.0790 | 1.9706 |
| **Referral Source - Other Clinical Specialty** | 0.3183 | 0.0753 | -0.0325 | 0.6691 |
| **Marital Status - Not Known** | 0.4639 | 0.0821 | -0.0590 | 0.9867 |

Service users incorrectly predicted to have high-intensity use (FP) were more likely to be Asian, to live with partners, children, or other relatives, and to have been discharged after the first assessment. Conversely, Black and mixed-race individuals, those referred from the voluntary sector, by carers, or via probation services, were more likely to be correctly identified as high-intensity service users by the original model.

### W1 (first assessment to the 3-month prediction point)

Table 4 illustrates the features at the end of W1 that were associated with an FP prediction, together with their corresponding coefficients. FPs were less likely among individuals whose initial screening had indicated significant risk or the need for further investigation, those on typical or atypical antipsychotic medications, patients with a recorded GP, and those actively engaged with multiple professional contacts, particularly through nurse and care coordinators. On the other hand, FPs were more likely in those with higher community treatment days, frequent address changes, outpatient status at the end of W1, and higher numbers of crisis attendances. A range of NLP-derived symptoms were also included within this analysis, at the end of both W1 and W2. FP prediction was more likely in patients with mood instability and less likely in those with early morning waking or disorganised symptoms by the end of W1.

**Table 4:** logistic regression results showing features at the end of W1 associated with a false positive prediction, with corresponding coefficients, p-values, and 90% confidence intervals.

| **<u>Variable</u>** | **<u>Coefficient</u>** | **<u>P value</u>** | **<u>90% confidence interval</u>** |  |
| --- | --- | --- | --- | --- |
| **Oral atypical antipsychotics** | -0.5073 | 0.0000 | -0.7282 | -0.2865 |
| **Brief Risk Assessment Screening Result - 'Significant Risk'** | -0.6811 | 0.0002 | -1.0453 | -0.3169 |
| **Brief Risk Assessment Screening Result - 'Not Recorded'** | -0.3064 | 0.0011 | -0.4907 | -0.1221 |
| **Nurse Event Count** | -0.0179 | 0.0019 | -0.0292 | -0.0066 |
| **NLP Symptom - Early Morning Waking** | -0.2918 | 0.0037 | -0.4887 | -0.0949 |
| **Accepted Team Count** | -0.1134 | 0.0042 | -0.1911 | -0.0358 |
| **Brief Risk Assessment Screening Result - 'Need for Further Investigation/Referral'** | -0.2970 | 0.0048 | -0.5035 | -0.0904 |
| **Care Coordinator Count** | -0.1016 | 0.0360 | -0.1965 | -0.0067 |
| **Typical antipsychotics - long-acting injectables** | -0.4624 | 0.0636 | -0.9510 | 0.0261 |
| **GP Recorded?** | -0.2116 | 0.0804 | -0.4487 | 0.0256 |
| **NLP Symptom - Disorganised Symptoms** | -0.0588 | 0.0816 | -0.1249 | 0.0074 |
| **Status at the end of W1 - Outpatient** | 2.8898 | 0.0000 | 2.4037 | 3.3759 |
| **Crisis Attendances** | 0.2775 | 0.0025 | 0.0979 | 0.4572 |
| **Address Change** | 0.2675 | 0.0153 | 0.0514 | 0.4835 |
| **NLP Symptom - Mood Instability** | 0.0285 | 0.0507 | -0.0001 | 0.0570 |
| **Days in Contact with Home Treatment Team** | 0.0057 | 0.0613 | -0.0003 | 0.0118 |

### W2 (3-months post prediction)

W2 prediction modifiers reflect measures derived from the immediate three months following prediction. Table 5 outlines the factors associated with an FP prediction after this point, and their associated coefficients. FPs were less likely among those who remained outpatients at the end of W2, experienced higher inpatient days, were under the care of multiple teams, and receiving input from social worker, consultant, or clinical psychologist. Additionally, a recorded diagnosis of personality disorder during W2 reduced the likelihood of an FP prediction, although the effect size was small. FPs were more common among patients with recorded use of cannabis or cocaine, receiving antidepressants or anticonvulsants, experiencing address changes, and those with higher numbers of crisis attendances. Input from forensic psychiatry was also associated with FP status. FP status was less likely in patients with recorded derailment of speech, guilt, or anxiety symptoms, and more likely in those with recorded poor appetite and thought withdrawal.

**Table 5:** logistic regression results showing features at the end of W2 associated with a false positive prediction, with corresponding coefficients, p-values, and 90% confidence intervals.

| **<u>Variable</u>** | **<u>Coefficient</u>** | **<u>P value</u>** | **<u>90% confidence interval</u>** |  |
| --- | --- | --- | --- | --- |
| **Status at the end of W2 - Outpatient** | -1.0719 | 0.0000 | -1.3288 | -0.8151 |
| **Temporary Address count** | -0.5912 | 0.0000 | -0.8747 | -0.3076 |
| **HCA Event Count** | -0.3008 | 0.0000 | -0.3890 | -0.2127 |
| **OT Event Count** | -0.1627 | 0.0000 | -0.2268 | -0.0985 |
| **Social Worker Event Count** | -0.1559 | 0.0000 | -0.2254 | -0.0863 |
| **Clinical Psychology Event Count** | -0.1393 | 0.0000 | -0.1720 | -0.1067 |
| **NLP Symptom - Anxiety** | -0.0874 | 0.0000 | -0.1148 | -0.0601 |
| **Total Document Count** | -0.0500 | 0.0000 | -0.0580 | -0.0420 |
| **NLP Symptom - Weight Loss** | -0.2022 | 0.0003 | -0.3128 | -0.0916 |
| **Distinct Generic Medications** | -0.1509 | 0.0023 | -0.2478 | -0.0539 |
| **Accepted Team Count** | -0.2308 | 0.0026 | -0.3813 | -0.0803 |
| **Consultant Event Count** | -0.1146 | 0.0038 | -0.1923 | -0.0369 |
| **Inpatient Days at the end of W2** | -0.6180 | 0.0043 | -1.0416 | -0.1943 |
| **NLP Symptom - Guilt** | -0.1322 | 0.0152 | -0.2389 | -0.0255 |
| **Typical antipsychotics - long-acting injectables** | -0.7500 | 0.0301 | -1.4278 | -0.0723 |
| **Personality Disorder Diagnosis at W2** | -0.3483 | 0.0896 | -0.7505 | 0.0539 |
| **NLP Symptom - Derailment of Speech** | -0.8078 | 0.0900 | -1.7415 | 0.1260 |
| **Address Change** | 0.4098 | 0.0037 | 0.1331 | 0.6864 |
| **Anticonvulsant Usage** | 0.5284 | 0.0090 | 0.1318 | 0.9250 |
| **Crisis Attendances** | 0.5838 | 0.0094 | 0.1431 | 1.0246 |
| **Cannabis Usage** | 0.0341 | 0.0201 | 0.0053 | 0.0628 |
| **NLP Symptom - Poor Appetite** | 0.2307 | 0.0209 | 0.0349 | 0.4264 |
| **Antidepressant Usage** | 0.2572 | 0.0215 | 0.0380 | 0.4765 |
| **Cocaine Usage** | 0.0581 | 0.0514 | -0.0004 | 0.1166 |
| **NLP Symptom - Thought Withdrawal** | 0.7452 | 0.0618 | -0.0366 | 1.5270 |
| **Other Alert Recorded** | 0.3671 | 0.0697 | -0.0296 | 0.7638 |
| **Consultant Specialty - Forensic Psychiatry** | 2.8277 | 0.0725 | -0.2581 | 5.9134 |
| **Lithium Usage** | 0.5640 | 0.0769 | -0.0610 | 1.1890 |

## Discussion

This study evaluated the performance of a previously validated logistic regression model for predicting high-intensity service use among 46,183 mental health service users assessed between 2007 and 2024. Of the 21,818 service users predicted to require high-intensity care, 4,174 were TP and 17,644 were FP Factors associated with FP predictions were examined over three timeframes: baseline (demographics, referral source, initial assessment), W1 (the 3-months post-first assessment), and W2 (the 3-months post-prediction). In interpreting the findings, it is important to distinguish between modifiers measured at baseline and during W1 compared to those from W2. Baseline and W1 factors inform the likelihood that a positive prediction is accurate at the time it is made, effectively providing a measure of confidence in the model’s output. However, W2 factors reflect events occurring after the prediction and potentially highlight circumstances or interventions that may alter eventual service use.

### Referral Source

The observation that referrals from socially engaged sources (e.g. voluntary sector organisations, carers, and probation services) were more likely to receive TP predictions has important implications for mental health service configuration. These referral sources may possess deeper knowledge of their users’ clinical conditions and social circumstances, arising from their ongoing relationships with clients and their position within community support networks, enabling them to predict with greater confidence when someone is likely to become unwell and require intensive services. The voluntary sector’s role in advocating for service users and their ability to engage individuals who may be reluctant to disclose mental health issues to statutory services makes them particularly valuable referral sources. Similarly, probation services work daily with individuals with high rates of mental health problems, and practitioners within these services have expertise in identifying likely cases of mental illness^6^. In contrast, emergency department referrals showed lower prediction accuracy from the original model, likely reflecting the transient nature of these encounters and the acute crisis-focused assessment environment, which may not capture the full complexity of ongoing mental health needs.

### Family Structure and Living Arrangements

Positive predictions among individuals living with partners, children, or other relatives were more likely to be false, which may reflect changing nature of these circumstances following the prediction point. For example, family members may become more involved once concerns are identified, helping to stabilise the person’s condition and avert the need for intensive care, thereby rendering the original high-intensity prediction incorrect^7^. Similarly, the finding that being widowed or a surviving civil partner was associated with false positive predictions may relate to the transient nature of bereavement-related stress. Individuals who experience recent loss may initially present with higher risk, but as they adjust over time, their likelihood of requiring intensive services may decrease, leading to apparent overprediction. It is also worth noting that marital status data had substantial missingness. The presence of any recorded information might indicate closer clinical engagement, meaning these cases were more actively managed and therefore more likely to deviate from their original predicted trajectories.

### Ethnic Disparities

The ethnic group variations in prediction accuracy may reflect well-documented disparities in mental health service access and utilisation. High-intensity service use predictions for Black or Black British service users were more likely to be correct, suggesting that the algorithm performs more accurately within this group. This is consistent with established literature showing higher rates of involuntary pathways into care, greater likelihood of crisis presentations, and generally poorer mental health outcomes for this population. These disparities have been previously described as potentially reflecting the cumulative impact of systemic racism, social disadvantage, and barriers accessing early intervention services^8^. Conversely, predictions for Asian and Bangladeshi service users were more likely to be FPs, indicating that the model was less accurate for these groups. One possible explanation is that family or community support, or changes in engagement after the prediction point, may buffer against deterioration and reduce the need for intensive care, leading to overprediction (as described earlier). Cultural factors, such as stigma around mental health or variations in help-seeking behaviour, may also contribute to lower recorded service use relative to predicted need^9^. These findings suggest that predictive models may require further adjustment to account for social and cultural contexts influencing service engagement.

### Antipsychotic Medications

The higher prediction accuracy observed among individuals prescribed antipsychotic medication may reflect greater availability of clinical information within this group to inform the original prediction model. People receiving depot or oral antipsychotics tend to have more frequent service contact, more detailed clinical records, and clearer diagnostic formulations, which may enhance the model’s ability to predict future service use accurately^10^.

### Patterns of Service Use

The association between service engagement and prediction accuracy likely reflects differences in the availability and consistency of clinical information between groups of service users that are more or less engaged. Individuals with higher numbers of recorded GP contacts, care coordinator interactions, or nurse events tended to have more accurately predicted outcomes. This may be because their ongoing engagement generates richer, more structured data and more consistent patterns of care, which the model can detect and extrapolate from effectively.

Crisis attendances appear to be associated with false positive predictions at both W1 and W2. When an individual has multiple crisis attendances leading up to the prediction, the model is more likely to overestimate the likelihood of future high-intensity service use. Furthermore, ongoing crisis attendances after the prediction continue to be linked with reduced prediction accuracy at twelve months. Clinically, this may reflect crisis services providing timely, targeted interventions that prevent escalation to longer-term intensive care. In practice, frequent crisis attendance could indicate that acute needs are being addressed effectively in the short term, buffering the progression to high-intensity service use.

The temporal patterns in outpatient status provide additional insights. Being an outpatient at the end of W1 was associated with FP predictions. Individuals predicted to require high-intensity care were often still receiving only low-intensity outpatient services, resulting in the model overestimating their eventual service use. By contrast, remaining an outpatient during W2 was associated with TP predictions. Continued outpatient engagement post-prediction may reflect persistent or escalating healthcare needs that signal a trajectory toward higher-intensity service use over the longer term. Taken together, this may suggest that early outpatient status could temper the accuracy of short-term predictions, whereas sustained outpatient engagement provides additional information that reinforces the accuracy of the model’s prediction.

### Markers of Care Complexity and Service Challenges

The emergence of certain clinical markers during W2 (e.g. assigned personality disorder diagnoses) was associated with TP predictions of high-intensity service use. This may reflect individuals with complex care needs who require sustained engagement with mental health services. Such markers can indicate a combination of factors, including higher comorbidity, challenging psychosocial circumstances, and patterns of service utilisation that strain standard outpatient management^11^. The observed association may also partly reflect systemic factors, such as service withdrawal or strained patient-staff relationships following the diagnosis, rather than the diagnosis itself inherently driving higher-intensity care.

### Strengths and Limitations

This study has several notable strengths. By leveraging a sample of over 46,000 patients across a 17-year period, it provides robust statistical power and the ability to capture long-term, naturalistic patterns in service use. The dataset represents near-complete catchment coverage, enhancing the generalisability of findings within this healthcare setting. Moreover, it comprised highly granular clinical, demographic, and service-use data, supplemented by natural language processing (NLP)–derived metadata, which allowed for a detailed and nuanced examination of factors influencing model predictions.

A limitation is that the original prediction model under evaluation uses observed healthcare costs and service contacts as proxies for clinical need, which may not fully capture true mental health acuity. The model assumes that individuals classified in the top 10% of predicted risk represent the highest users of services, implying that high-intensity service use is generally observable and not ‘hidden’. However, this assumption may not hold for all individuals. Some people with significant mental health needs may not progress to high-intensity service use due to barriers such as stigma, cultural appropriateness of services, or trust in healthcare providers. Additionally, out-migration from the catchment area may result in underestimated service use, leading to apparent FP predictions. Finally, certain factors that potentially influence mental health trajectories, including social stressors, experiences of discrimination, or levels of informal support, were not available in the dataset, which will have reduced model accuracy. Consequently, some FP classifications may reflect limitations in observed service data or unmeasured risk factors, rather than a true mismatch between predicted and actual clinical need.

The analysis utilises data from a single catchment-based mental health service, representing a limitation for model generalisability and external validity. Single-site studies are vulnerable to institution-specific practices, patient demographics, service configurations, and local resource constraints that may not be representative of other healthcare settings. On the other hand, the source service serves a diverse patient population across a wide range of services within a defined catchment area, providing comprehensive coverage that mitigates some concerns about homogeneity. Nevertheless, external validation across multiple sites is essential to demonstrate model transportability and assess performance in different settings.

## Conclusion

This workflow could potentially be implemented into clinical practice in a 2-step manner, wherein the earlier model by Chada, Stewart and Lai is first applied to identify high-risk service users, and this second model is used to determine the ‘believability’ of the earlier model’s predictions. Baseline and W1 data help determine how likely a positive prediction is to be correct at the time of prediction, while W2 factors can help explain why some initially predicted high-risk individuals do not progress to high-intensity service use. Regular monitoring and recalibration will be essential to maintain model performance over time, particularly given the dynamic nature of healthcare systems and evolving patient populations. Additionally, efforts should be made to address identified biases through improved data collection practices, enhanced access to services for under-represented populations, and the development of more equitable predictive models that account for social determinants of health and access barriers.

These findings have important implications for how predictive models in mental health could be interpreted and applied in clinical practice. The identification of factors (at baseline and during W1) associated with TP vs. FP predictions provides clinicians, care coordinators, and other model users with additional context for interpreting model outputs. Predictions may be considered more reliable in the presence of multiple TP predictors (i.e. socially engaged referral sources, antipsychotic medication use, sustained family disconnection), while caution may be warranted when FP predictors predominate (certain ethnic backgrounds with strong family support, crisis service engagement patterns). This approach supports a more nuanced application of predictive modelling that considers not just the primary prediction, but the myriad factors that may contribute to the accuracy of that prediction. Such contextualisation should enhance clinician confidence in model outputs and support more appropriate resource allocation decisions, ultimately improving both the efficiency and effectiveness of mental health service delivery.

## Data Availability

The data for this study were sourced from South London and Maudsley NHS Foundation Trust (SLaM) electronic health records through the Clinical Record Interactive Search (CRIS) platform. Metadata, including variables derived using natural language processing (NLP), were generated with the TextHunter text-mining software. All analyses were performed within a secure virtual desktop environment, with no patient data leaving this system. Access was restricted to authorised researchers holding the necessary approvals, such as a research passport and CRIS training. Data processing and analyses were conducted using Python and Microsoft Excel. Due to confidentiality and governance restrictions, the data are not publicly available; however, qualified researchers may obtain access through SLaM CRIS, subject to institutional approval and data access agreements.

## Notes

### Competing Interest Statement

The authors have declared no competing interest.

### Funding Statement

Funding Statement: RS is part-funded by: i) the NIHR Maudsley Biomedical Research Centre at the South London and Maudsley NHS Foundation Trust and Kings College London; ii) the National Institute for Health Research (NIHR) Applied Research Collaboration South London (NIHR ARC South London) at Kings College Hospital NHS Foundation Trust; iii) UKRI Medical Research Council through the DATAMIND HDR UK Mental Health Data Hub (MRC reference: MR/W014386); iv) the UK Prevention Research Partnership (Violence, Health and Society; MRVO49879/1), an initiative funded by UK Research and Innovation Councils, the Department of Health and Social Care (England) and the UK devolved administrations, and leading health research charities; v) the NIHR HealthTech Research Centre in Brain Health. This research was carried out by BVC while in receipt of funding from Kings College London, and as part of the NHS Fellowship in Clinical Artificial Intelligence (AI), funded by the NHS Digital Academy and Guys and St Thomas NHS Foundation Trust.

### Author Declarations

The CRIS Oversight Committee of the South London and Maudsley NHS Foundation Trust gave ethical approval for this work.

